# LLM-assisted Clinical Coding Audit through an Interpretable Coding Pipeline

**DOI:** 10.1101/2025.08.24.25334321

**Authors:** Supriya Khadka, Xiaorui Jiang, Vasile Palade

## Abstract

Clinical coding is a vital yet complex part of healthcare practice. Most automated coding research relies on imperfect training data, which has an unavoidable negative impact on the quality of code prediction. A key contributing issue, vastly overlooked in current research, is the ubiquitous presence of undercoding and various types of coding errors in widely used coding datasets. From another angle, coding audit is as challenging as coding itself due to the lack of assistive tools, which is also under-studied compared to automated coding research. In this work, we uncover substantial undercoding and errors in commonly used datasets and present the first empirical study on their impacts on the performances of automated coding algorithms. To enable this, we develop an interpretable coding pipeline, using large language models (LLMs) for evidence extraction and code verification, and a multiclass classifier trained on a large-scale dataset of silver-standard evidence-code pairs for code prediction. Assisted by the pipeline, three professional coders systematically identify, categorise, and correct errors in two widely-used coding datasets that have human-annotated evidence texts for assigned codes. As an AI-assisted coding audit tool, the current study uncovers significant data quality issues, including a 76.3% undercoding rate in MDACE and a 29.7% error rate in CodiEsp. Re-evaluating existing models on error-corrected datasets results in consistent performance improvements. Additionally, the pipeline is a highly potential coding framework, which achieves superior or comparative performances to state-of-the-art LLM-based methods. The results underscore the necessity of shifting research focus from model-centric to data-centric solutions in clinical AI.

## I. Introduction

CLINICAL coding is the process of assigning systematic codes that represent diagnoses, treatments, lab tests, and medications, etc. to patients’ medical records. Clinical coding is crucial for billing, insurance claims, policymaking, and healthcare analysis. In practice, clinical coding is mainly manually performed by professional coders, making the process time-consuming and error-prone [1]. In the past few decades, researchers have been working on developing automated clinical coding methods [2-3], and most of them were on International Classification of Diseases (ICD) [4-7], likely because of its universal adoption over decades and the availability of public datasets [8-9].

Many challenges exist for developing effective AI-based automatic clinical coding (ACC) solutions. Well-known issues include multiple codes per documents to be predicted from a large space (e.g., over 70K codes in the growing ICD-10 set), inadequate up-to-date public data of high code coverage, data sparsity for many codes that only appear a handful of times, lack of interpretability and trustworthiness of black-box deep learning-based models, etc. [10] Among these challenges, **code quality issues**—e.g., miscoding, undercoding/overcoding [11], and upcoding/downcoding [12]—have long been under-studied by the ACC community but reported as wide-spread in health practice [13] and impactful for health administration, billing and research [14]. Every year, coding errors cause a huge number of inaccurate payments and significant loss of financial renumeration to health providers [15].

Code quality and ACC performance are two intertwined factors. Existing studies of coding accuracy have found that a large portion of errors [16-18] are ubiquitously existent in human-assigned codes in clinical documents across all medical fields [19-25]. Because ACC models are trained on datasets that rely on human-assigned codes as gold standards [8-9], improving code quality obviously has a positive impact on ACC performance. However, these studies were all conducted by manual investigation. Although there have been many advancements in ACC based on deep learning and large language models (LLM) [6-7], they all focused on ACC models for code assignment but neglected their potentials for code quality analysis, coding audit and miscoding correction.

The current study proposes an LLM-assisted interpretable coding pipeline that “kills two birds with one stone”. While our focus is to identify and analyse the quality issues in existing ICD coding datasets, the proposed approach also serves as a promising framework for LLM-assisted clinical coding and the future research. In particular, this paper focuses on identifying two broad types of issues prevalent in existing ICD datasets: **undercoding** and (other types of) **coding errors**. Given that datasets differ across institutions, both in terms of text style and annotation strategy [26], we observed corresponding variations in the error types they contain. These differences also affect the performance of state-of-the-art (SoTA) ACC models.

In short summary, we designed a three-stage code prediction pipeline: an LLM-based evidence extractor, a multiclass classifier that is trained on the silver-standard evidence (*silver evidence* hereafter) obtained from the MIMIC-IV dataset [9], and an LLM-based verifier to produce the final set of codes. These codes were then analysed for undercoding and coding errors using two complementary approaches: **ensemble-based crosscheck** and **manual review**, as shown in Fig. 1

**Fig 1.**
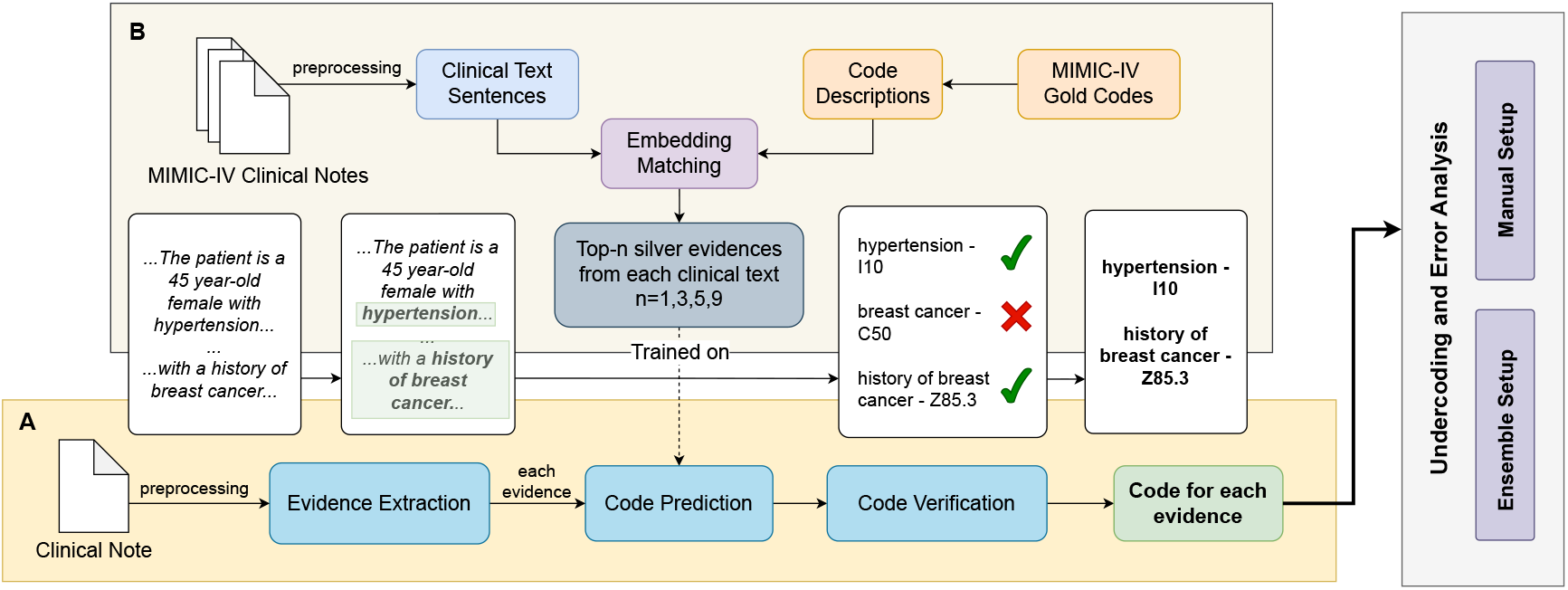
Overview of the proposed methodology. (A) Code prediction pipeline with evidence extraction, prediction, and verification modules. (B) Silver-evidence creation process used to train the predictor. (C) Undercoding and error analysis through ensemble and manual verification. Components with a white background highlight illustrative examples.

. Noticeable performance improvements were observed when re-evaluating SoTA deep learning-based ACC models on the error-corrected datasets. Meanwhile, our pipeline achieved outstanding ICD coding performances compared to the SoTA LLM-based ACC approaches. Our major contributions are:

- Identification and quantification of undercoding and (other types of) coding errors in widely used ICD coding datasets with human-annotated rationales;
- Demonstration of performance improvements in existing models when evaluated on corrected datasets;
- Proposal of a promising, interpretable clinical coding pipeline that synergises large and small language models to provide valuable diagnostic insights for code auditing and suggestion without relying on large-scale annotated datasets of coding rationales.

## II. Related Work

### A. Deep Learning for Clinical Coding

Automated clinical coding has predominantly been framed as a multi-label classification problem, as each clinical note may be assigned multiple ICD codes. Initial efforts employed traditional machine learning methods [3, 26, 28], and soon deep learning models based on architectures like Long-Short Term Memory (LSTM) [29-30] and Convolutional Neural Network (CNN) [31-32] became dominant, which were also often augmented with attention mechanisms [33-36]. Among recent methods, PLM-ICD [37] that is based on pretrained language models has achieved state-of-the-art results on many public coding datasets of MIMIC [8-9], as shown in a comprehensive reproducibility study [38].

### B. Evidence-based Clinical Coding

Interpretability of AI solutions is a key issue in the clinical setting. Several studies have attempted to solve this issue by identifying the evidence behind each code. Often attention mechanisms were used to highlight the text segments as code rationale or supporting evidence for predicted codes [39-40]. Alternatively, studies also claimed that unsupervised methods can also identify coding evidence and achieve explainability levels comparable to supervised models [41]. Along the line of explainable ACC, several ICD coding datasets with evidence have been created, such as CodiEsp in CLEF eHealth 2020 [42] (death certificates in Spanish), MDACE [43] (a subset based on MIMIC-III), RD-IV-10 [44] (a subset derived from MIMIC-IV), and ACI-Bench [45] (synthetically created doctor-patient conversations). Closest to our study is the silver-standard for MIMIC-III clinical coding dataset developed in [46].

### C. LLMs in Clinical Coding

There are a few recent attempts of benchmarking LLMs’ (zero-shot) capabilities for understanding clinical codes [47-49] and assigning codes [50-52]. Despite of inferior performances to deep learning methods, LLM has shown unique advantages in rare code prediction [53], domain transfer [54], and evidence extraction (current study). LLM-based ACC has attempted to address several aspects akin to human coding suggested in [55], such as employing the ICD code hierarchy (*aka*. tabular index) for step-by-step coding [56] and navigating the alphabetical index of the ICD guideline [57-58].

Recent studies have been focusing on evidence-based ACC. In [59], first codes are predicted and then evidence is extracted to verify each code prediction. Other studies either first extract evidence to provide the basis for reasoning for code prediction and justification [45, 57, 60-61] or perform evidence extraction and code prediction at the same time [58, 62-63]. Retrieval-augmented generation was also applied to strengthen LLMs’ reasoning abilities for coding by utilising semantically relevant contexts [57] and coding examples [58]. Methodologically, our work has made a unique contribution by proposing a novel interpretable clinical coding framework useful for both clinical coding and coding audit. Apart from [46], our work is the only study to enrich a large ICD coding dataset with high-quality silver evidence and train evidence-based code classifier.

### D. Coding Accuracy and its Impact

While ignored by most ACC studies, coding accuracy has long been a notable issue in health data management [13, 64] as inaccurate coding causes significant financial loss to the health service [15] and problems to health data administration [14]. Studies about coding accuracy are prevalent [16-18, 65-66] across many clinical subjects [19-25, 67-68]. The inaccuracy rates varied. Some studies reported very high inaccuracy rates (including both undercoding and other coding errors) over 55% [16, 69-71], even above 70% [18] or 80% [44] in some extreme cases. Similar to our own analysis, undercoding is a significant phenomenon, reaching 40-60% in many studies [20, 23-24, 67]. In most cases, high rates of undercoding and coding error cause huge loss in financial renumeration to health providers [16].

### E. Coding Aduit and Error Correction

Comparatively, studies about methods for code correction or coding audit using technology are relatively sparse. Existing studies are mainly based on data mining [72-73], such as association rule mining for co-existent codes [74-75]. Only a few studies trained classifiers to detect code assignment error [76-77], including refining unspecified condition codes [78]. Like our analysis, widespread undercoding in MIMIC-III was detected in [46] particularly among frequent codes, with similar findings reported in [79]. To the best of our knowledge, there does not seem to be any work on using AI to assist large-scale coding audit and quality analysis.

## III. Code Prediction Pipeline

We found the most effective way to identify undercoding and coding errors was to build a code prediction pipeline that offers transparent explanations at each step, clarifying why a code is included or omitted. The pipeline starts with clinical text preprocessing, followed by mapping extracted evidence spans to ICD-10 codes. Finally, an LLM-based verifier reviews the full text to filter out any inappropriate codes.

### A. Clinical Text Preprocessing

We used a variety of preprocessing strategies depending on the nature of clinical texts. Datasets based on MIMIC (Medical Information Mart for Intensive Care) [43, 54] contain longer, messier, but more structurally consistent notes (e.g., discharge summaries). In addition to the information-rich sections such as *Chief Complaint, History of Present Illness, Past Medical History, Family History, Social History*, and *Discharge Diagnosis* [80], we also found human-annotated evidence in other sections so they were also extracted: *Medications, Brief Hospital Course*, and *descriptions of the patient’s critical care status*. Many extracted phrases contained non-clinical or coding-irrelevant details, so they were passed through an evidence extraction module, which is designed to retain only clinically meaningful codable information (Sect. II.B).

Often, a sentence may contain medical evidence that justify multiple codes, especially when different conditions are listed together. To isolate distinct clinical concepts, we split sentences into smaller phrases using punctuations (e.g., commas, colons, semicolons) as well as conjunctions (like “and”). Unnecessary punctuation and de-identified content were removed from these phrases. Each word from the phrases is then passed through a manually maintained lookup table to expand abbreviations, which can be found at: GitHub link after being accepted.

In contrast, as CodiEsp is structurally simpler and sentences are often densely packed with clinical information, it requires less preprocessing. The same phrase-splitting preprocessing using punctuation- and conjunction-based rules was applied. As the original clinical texts in CodiEsp are in Spanish and the English translations provided have very poor qualities, often omitting critical clinical details present in the original Spanish texts, we re-translated them using Google Translate in our analysis. Also note that given the smaller number of phrases and the sentence-level structure, we chose to omit the evidence extraction step for the experiments on CodiEsp.

### B. Evidence Extraction

The purpose of the evidence extraction module is to ensure retaining only clinically relevant and codable phrases. Inspired by [59, 62], evidence extraction was accomplished using LLMs. Fig. 2 shows the prompt. It was initially designed in a simple way to include *any medical condition, symptom, diagnosis, finding, or infection resulting from a procedure* (see the “Include sentences if it contains” section). During tuning the prompt, we observed that since LLMs sometimes extracted content that was clinical in nature but not relevant for coding, such as lab results, negative diagnoses, or general narrative text, and occasionally missed important codable details, such as causes of condition, patient’s social history, or critical care information. Therefore, we iteratively enriched the prompt by introducing clinically meaningful “exclusion rules” (see the “Exclude sentences if it contains” section). Initial experiments also showed that few-shot approach (2–3 examples for each clause in our experiments) worked best for evidence extraction, particularly in guiding what to include and exclude.

**Fig 2.**
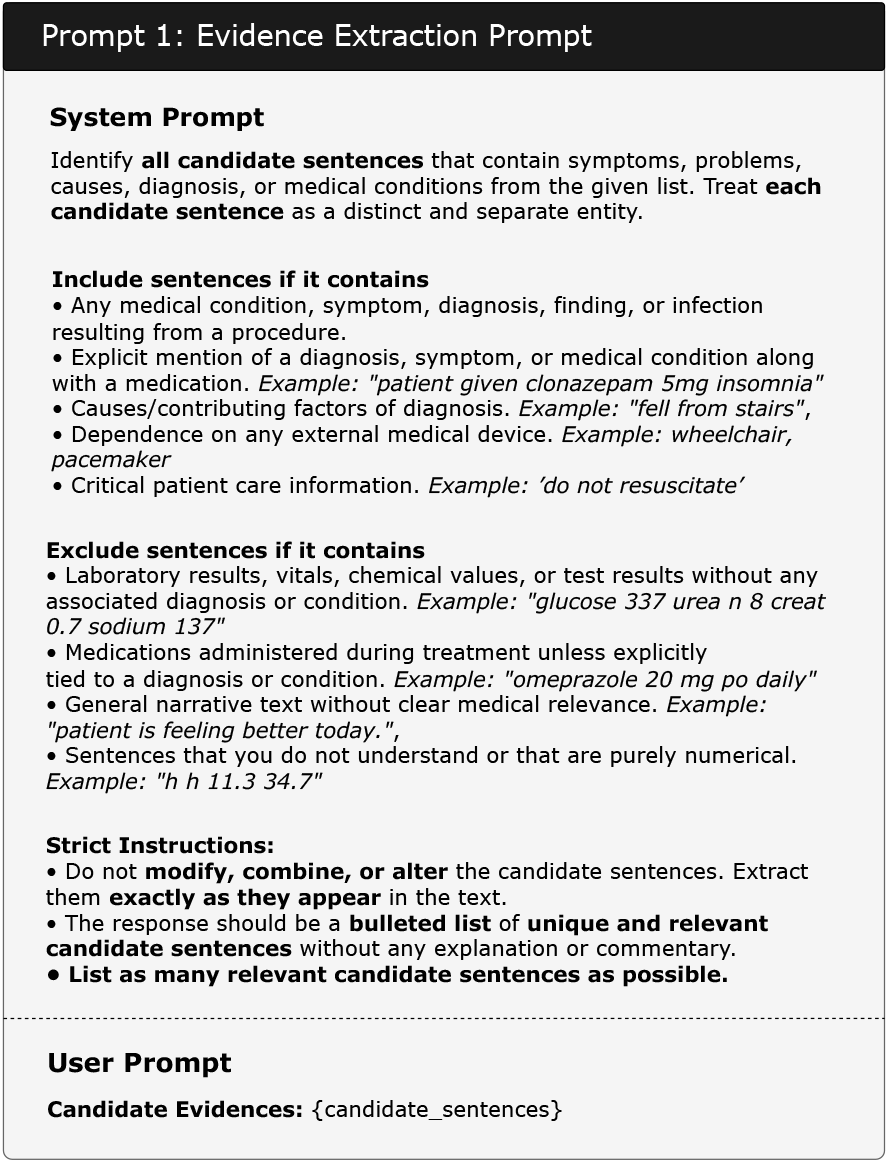
Prompt for Evidence Extraction.

### C. Code Prediction

Codes are then predicted based on the extracted evidence. To accomplish this, a multiclass classifier is trained to assign a code to a given evidence phrase. Because a large-scale dataset suitable for training such a classifier does not exist, one of our major contributions is to construct a custom dataset of silver evidence from MIMIC-IV, where each entry contains a human-assigned ICD-10 code and a corresponding evidence phrase.

#### 1) Silver Evidence Dataset Creation

The dataset creation process is illustrated in Part (B) of Fig. 1. We first preprocess the clinical texts of MIMIC-IV following the same procedure described in Section II.A. In parallel, for each clinical note, we collect descriptions of the “gold-standard” human-annotated ICD codes and map each code description to the most relevant evidence phrase(s). The mapping was done using the Sentence-BERT model MiniLM [81]. Although the most relevant top few phrases can be retrieved for each code description, in this paper the top match is selected as a silver evidence instance to create the silver evidence dataset for the sake of simplicity. LLM-based evidence extraction was not employed for silver evidence extraction due to the scale of MIMIC-IV (over 10K long texts) and LLMs’ tendency of over-extraction. In our additional experiment, a SoTA model PLM-ICD [37] trained on MIMIC-IV using top-*n* matched evidence phrases achieved relative improvements of over 20% when *n* ≥ 3. This demonstrates the meaningfulness of silver evidence (Table VIII in Appendix).

Further validation was performed by evaluating this method against human-annotated evidence spans in MDACE. For each code in MDACE, we extracted the silver evidence phrases and assessed the resulting matches. Our method achieved an 85% match rate between the extracted and annotated evidence. Most mismatches occurred because our method identified a piece of text that better aligned with the code description than the original annotated evidence according to human review (Sect. IV.B). For example, for code I130, which corresponds to *Hypertensive heart and chronic kidney disease with heart failure and stage 1 through stage 4 chronic kidney disease, or unspecified chronic kidney disease*, MDACE has labeled “Hypertension” as the supporting evidence, but our method ranked “chronic systolic heart failure” and “chronic renal insufficiency baseline creat 1.5” as better matches, with “Hypertension” placed as the third-best match. “Hypertension” and “chronic systolic heart failure” combined arguably provide the most accurate clinical context for this code. But since the current version of our method extracts and evaluates individual phrases in isolation, such combinations can’t be captured so we conservatively classified this outcome as an inaccurate match.

#### 2) Training Code Classifier

A multiclass classifier is trained on the silver evidence dataset. Each ICD code is treated as a distinct class and each pair of code and the corresponding silver evidence as a training example. Prior to training, further preprocessing is needed. Since certain evidence phrases appear under multiple codes during the mapping procedure, we resolve these conflicts by selecting the code with the best-matching description based on embedding similarity. For example, while “chronic systolic heart failure” may appear as evidence for I130 (see the example above), it is more accurately aligned with I5022–*Chronic systolic (congestive) heart failure*. In such cases when a clinical note contains both codes, we use MiniLM embeddings to match the phrase to the most appropriate code.

We fine-tune ClinicalBERT [82] to train the code classifier. In the current version of our implementation, codes with fewer than five silver evidence examples are excluded from training to ensure meaningful representation. In total, we obtained silver evidence for 14,021 code instances, covering 6,311 unique codes. Many of the excluded infrequent codes had only a single instance in the training set and none in the test set, which made it impossible to evaluate performance reliably. On the one hand, we argue that including such codes may led to unstable training and misleading metrics, as the model either overfit to rare examples or failed to learn any meaningful pattern. By focusing on sufficiently represented codes (≥5 times in the current paper), we ensure better generalization and more robust classifier performance. On the other hand, we plan to resolve such few-shot or even zero-shot cases in our future work as in [83] and expect datasets with better code coverage.

### D. Code Verification

Inspired by recent attempts [54], the codes predicted on the extracted evidence are passed through an LLM-based verifier. This verification step aims to ensure filtering out any code that should not have been assigned, for example false positives arising from symptoms that are already covered by a diagnosis or from negative statements that may have been incorrectly split (e.g., “headache or nausea” parsed from “no fever, headache or nausea”, missing the restrictive modifier “no”).

We tested multiple prompts and input formats and optimised the code verification prompt as shown in Fig. 3. The predicted codes along with the original clinical text are feed as input to the code verifier. The simplest prompt, asking a LLM to extract supporting evidence if any exists for a given code, achieved the highest recall but suffered from low precision. In contrast, a more refined prompt that instructs LLMs to remove redundant codes or those already implied by other diagnoses yielded slightly lower recall but significantly higher precision.

**Fig 3.**
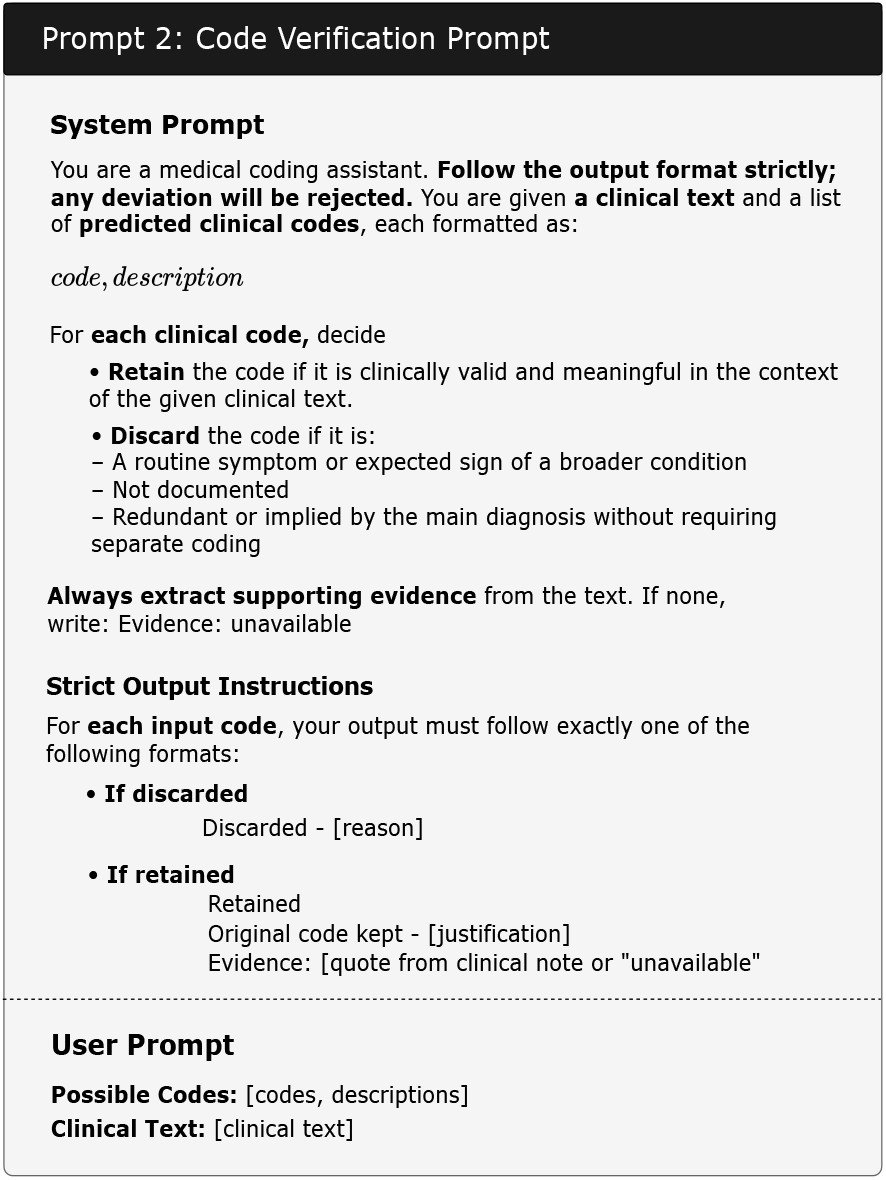
Prompt for Code Verification.

## IV. Undercoding and Error Discovery

**Undercoding** is a situation where codes that should have been assigned are not assigned. It is paid specific attention to because undercoding is one major source of potential renumeration loss. We use the term **error** to refer to any other form of coding issue observed in the datasets. We further classify errors into eight specific categories, as detailed in Table I in the results section.

**TABLE I.** Error Categories, Examples, and Proportions(among all Miscoding)

| Category | Definition | Example | MDACE(%) | CodiEsp(%) |
| --- | --- | --- | --- | --- |
| **Over-generalization** | Specific details missed; broader code used | *Right eye blindness* coded as *blindness, one eye, unspecified* | 14.23 | 1.46 |
| **Clinical Detail Mismatch** | Code misses key modifiers | *Coronary artery disease (CAD) with chest pain* coded as *CAD without chest pain* | 32.12 | 11.25 |
| **Contradictory** | Conflicting codes used together | *Infectious and noninfectious gastroenteritis* coded for the same text | 0.73 | 0.34 |
| **History as Active** | Past conditions coded as current | *History of breast cancer* coded as *active*. | 14.96 | 4.62 |
| **False Positive** | Code assigned despite negative results | *Urinary tract infection* coded despite negative findings | 1.46 | 20.95 |
| **No Evidence** | No clinical basis in text for the given code | *Hyperkalemia* coded with only *hyperlipidemia* as evidence | 5.11 | 10.53 |
| **Signs/Symptoms** | Symptom codes assigned despite more specific diagnosis | *Localized swelling* coded when *cellulitis* documented | 12.4 | 24.26 |
| **General Overcoding** | Codes added without clear clinical justification | *Depression* coded from brief low mood mention | 18.25 | 25.6 |

Two independent setups are used to identify undercoding and errors. In an **ensemble crosscheck** setup (detailed in Sect. IV.A), we aggregate the outputs from various combinations of LLMs acting as extractors and verifiers and flag probable errors using a strict two-thirds majority voting rule. In the **manual review** setup (detailed in Sect. IV.B), the responses (predicted codes and reasoning) from three best-performing LLM-based (extractor, verifier) pairs are assessed by three experienced human annotators against the original codes to identify undercoding and coding errors.

### A. Ensemble Crosscheck Setup

Let ℰ = {*E*_1_, *E*_2_,…, *E*_*m*_} and 𝒱 = {*V*_1_, *V*_2_, …, *V*_*n*_} denote the sets of LLM-based evidence extractors and code verifiers, respectively. Each extractor-verifier pair *M*_*ij*_ = *E*_*i*_ ∘ *V*_*j*_ defines a unique model combination, where 1 ≤ *i* ≤ *m* and 1 ≤ *j* ≤ *n*. Let ℳ = {*M*_*ij*_} represent the complete set of extractor-verifier models, with |ℳ| = *m* × *n*. Finally, let 𝒯 = {*t*_1_, *t*_2_, …, *t*_*T*_} be the set of clinical texts. For each clinical text *t* ∈ 𝒯, we denote:

- *K*_*t*_ ⊆ 𝒴: the set of gold-standard ICD-10 codes (i.e., the original codes assigned by professional coders), where 𝒴 is the universe of all possible codes;
- *P*_*ij*_(*t*) ⊆ 𝒴 : the set of ICD-10 codes predicted by model *M*_*ij*_ ∈ *M* for clinical text *t*.

We define the vote count for a code *y* ∈ 𝒴 as the number of model combinations that predict *y* for a given text *t* ∈ 𝒯:

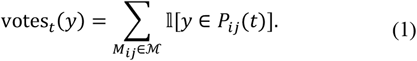

A code is included in the ensemble prediction set 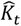 if it is predicted by at least two-thirds majority of the models:

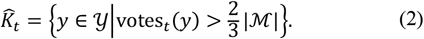

Based on (2), we define the following sets of (i) **True Positives** 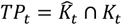, (ii) **Probable Undercoded Codes** 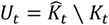, and (iii) **Probable Error Codes** 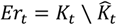, which is also evaluated against manual review (see Sect. VI.D).

### B. Manual Review Setup

All model combinations are evaluated on MDACE. As it is unscalable to manually annotate outputs from all models, the three best-performing combinations are selected for manual review (Fig. 4). Selecting only three LLM combinations may introduce bias. To mitigate this, the ensemble-based method is introduced in parallel, allowing both methods to complement each other. Each note is reviewed and re-annotated based on LLM-extracted evidence, along with the original clinical text and the logic used by LLMs to accept or reject specific codes, thereby supporting human reviewers in decision making. Codes predicted by any LLM combination are included for annotation.

**Fig 4.**
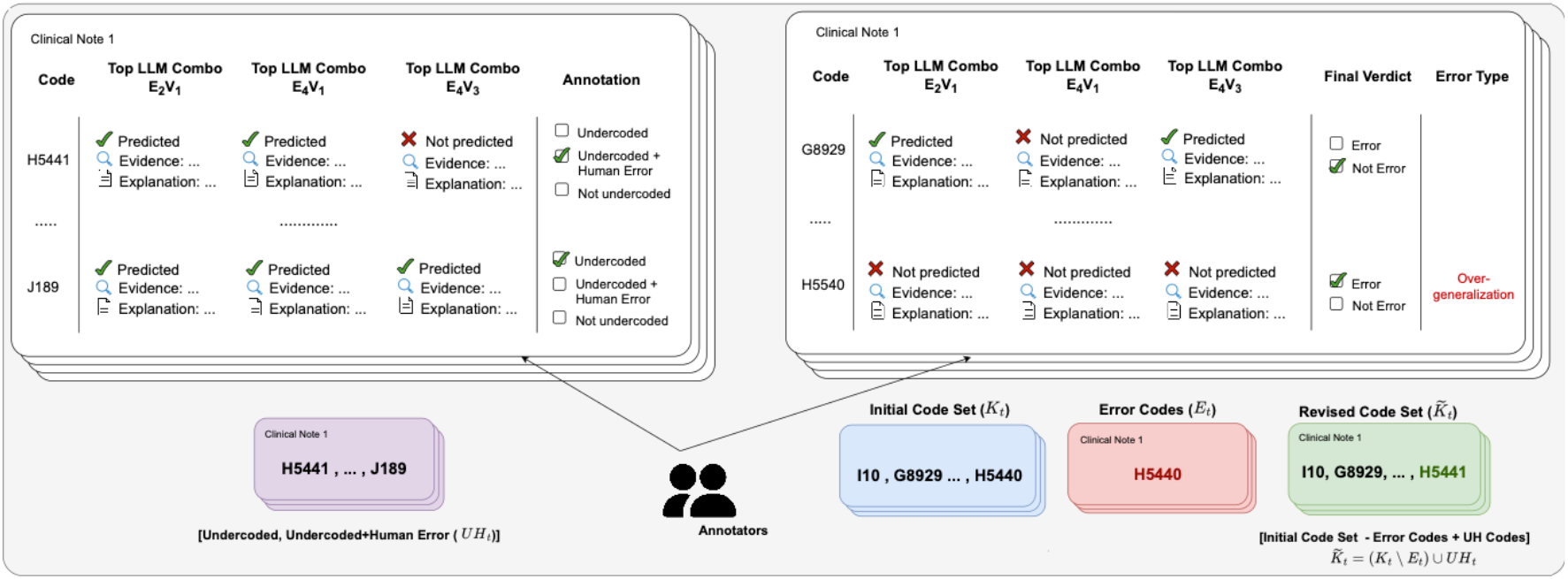
Manual Review Setup of Undercoding and Error Analysis.

Review was conducted by three skilled professional coders of in total over 20 years of experience. A sample of 10% notes was used for initial annotation by all three human reviewers to develop the (initial) error typology and measure inter-annotator agreement (IAA). Since very high agreement was achieved, the remaining documents were divide d among the reviewers. Error categories were iteratively refined and finalised in consultation with all three reviewers during the reviewing process.

For the manual review of code quality issues, we selected the best-performing model combinations on MDACE based on F1 scores (see Table V in the results section). To avoid bias from relying solely on one model, (GPT-4.1, GPT-4.1), (Gemini-2.0-Flash, GPT-4.1) and (Gemini-2.0-Flash, Gemini-1.5-Pro) were chosen to assist coding audit.

For undercoding analysis, each predicted code is annotated to one of the following three categories: (ii) an **Undercoded Code with Human Error** (*UH*_*t*_) is a undercoded code that is also the correct replacement for an erroneously assigned code in the original annotation; (ii) an **Undercoded New Code** (*UN*_*t*_) is a correct code based on the available evidence that is missed by the original annotation; (iii) a **Non-Undercoded Code** is a code that is incorrectly predicted by our pipeline, for which the evidence is insufficient to justify its assignment.

For error analysis, each original code is labeled as either an **Error** (*E*_*t*_), i.e., an original code that is incorrectly assigned based on the evidence, or a **Non-Error**, meaning the original code assignment is justified by the evidence. Errors are further categorized into one of eight error types, as detailed in Table I. Note that the last four error categories in Table I can be grouped under the broader category of *Overcoding*. However, we found sufficient examples to justify separate categories for *False Positive, No Evidence*, and *Signs and Symptoms*. The remaining and often isolated instances of other kinds of overcoding were grouped under a single category termed *General Overcoding*.

### C. Inter-annotator Agreement

To evaluate the overall consistency in error and undercoding detection, **binary agreement** was calculated, where reviewers determined whether a predicted code is undercoded or not and whether an originally annotated code is an error or not. For evaluating the reliability of finer-grained error categorization, we calculated **multi-class agreement**. For both cases, we used Krippendorff ‘s Alpha (*α*) for inte-annotator agreement (IAA) evaluation because it generalises several IAA metrics and can accommodate any number of annotators. In explorative studies, values above 0.8 are considered highly reliable, and values between 0.6 and 0.8 are seen as acceptable.

### D. Correcting Errors

As described in Section IV.B, we performed two kinds of annotation for each clinical text *t*: the set of coding errors *E*_*t*_ that were incorrectly assigned; the set of undercoded codes as replacements for some codes erroneously assigned by humans *UH*_*t*_. Based on them, we revised the gold-standard code set for each clinical text *t*, by replacing the identified error codes with their appropriate undercoded alternatives, formally as follows:

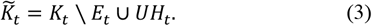

Equation (3) ensures that ACC models’ performances are more accurately reflected when evaluated against the revised gold standard set 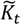, often with underreported improvements.

## V. Experimental Setup

### 1) LLMs

We experimented with 4 LLMs for both evidence extraction and code verification: GPT-4.1-mini, GPT-4.1, Gemini-1.5-pro, Gemini-2.0-flash, and Deepseek-R1. This resulted in a total of 16 extractor-verifier combinations (4 extractors × 4 verifiers). We followed the PhysioNet guidelines to use Azure to access GPT and Deepseek models, and Google Cloud Platform for Gemini.

### 2) Datasets

We mainly used two datasets that have human-annotated rationales for ICD-10 diagnostic codes: MDACE [43] and CodiEsp [42]. For MDACE, we used the in-patient “Discharge Summary” notes. After excluding addendums and the validation set, 239 clinical notes were included in our experiments. For CodiEsp, we conducted experiments on its test set, which contains 250 clinical texts.

### 3) Baseline Models for ICD Coding

We used four popular ACC models as baselines: CAML [39], MultiResCNN [31], LAAT [33] and PLM-ICD [37]. For MDACE, the baselines were trained using MIMIC-IV. As MDACE is a curated subset of MIMIC-III, the experiments to some extent evaluated cross-dataset generalisability. Due to significant domain mismatch and text style difference, these MIMIC-IV-trained models performed poorly on CodiEsp, so we created a hybrid training set by merging the reviewed parts of CodiEsp with the subset of MIMIC-IV that predominantly contain the codes founds in CodiEsp. This approach helped minimise the introduction of additional code complexity and allowed us to study how the erroneous nature of CodiEsp affects model performance.

## VI. Results

### A. Inter-Annotator Agreement is High

Before presenting findings about the code quality issues, we first report that very high IAA was achieved by the professional coders that we recruited on both undercoding and coding errors.

#### 1) Binary Agreement

For undercoding, the primary goal is to determine whether a prediction identifies a missing code. The resulting Krippendorff’s Alpha was **0.864**, considered as highly reliable. For coding errors, the binary evaluation (Error vs. Non-Error) resulted in a Krippendorff’s Alpha of **0.768**, lying in the upper range of tentative reliability and indicating reasonably high consistency across annotators.

#### 2) Multiclass Agreement

Next, we assessed the agreement across the eight specific error categories defined in Table I. For this analysis, we included only those codes where a majority of annotators agreed that an error was present. Majority agreement provides a balance between overly strict or lenient criteria— requiring unanimous agreement may exclude genuine errors if one annotator overlooks them, while accepting single-annotator judgments risks including spurious errors due to individual bias.

Some error types were frequent such as Clinical Detail Mismatch and Over-generalization, while others occurred much less often such as No Evidence. Despite this imbalance, we obtained an extremely high IAA, with a Krippendorff’s Alpha of **0.913**. We attribute this strong agreement to the fine-grained categorisation scheme, which provided clear distinction and reduced ambiguity for our reviewers to determine error types. This high multi-class reliability also helped offset the slightly lower binary error agreement, giving us confidence in the robustness of our code review process and reliability of the results of analyses in the following subsections.

### B. Undercoding Findings

In the ensemble setup, the three top-performing (Extractor, Verifier) combinations were: (GPT-4.1, GPT-4.1), (Gemini-2.0-Flash, GPT-4.1) and (Gemini-2.0-Flash, Gemini-1.5-Pro). See Sect. VI.E for details about the performance of the proposed code prediction pipeline. We found the undercoding rate on MDACE to be about **71.63%**. The most frequently occurring codes tend to be both undercoded and error-prone. In particular, Chapter Z codes (about “Factors Influencing Health Status and Contact with Health Services”) were among the most undercoded. We guess this is partly due to their high frequency in the dataset and partially because Chapter Z codes are not direct diagnoses so that they are often overlooked by coders.

In the manual setup, the (potential) undercoding rate on MDACE was as high as **76.34%**, showing a close alignment with the ensemble-based estimate. Both methods revealed similar trends in the most frequently undercoded codes, as shown in Fig. 5. There is a clear relationship between the actual frequency of a code and its likelihood of being undercoded, in both manual and ensemble settings. This shows that ensemble analysis can be more or less reliable in detecting undercoding.

**Fig 5.**
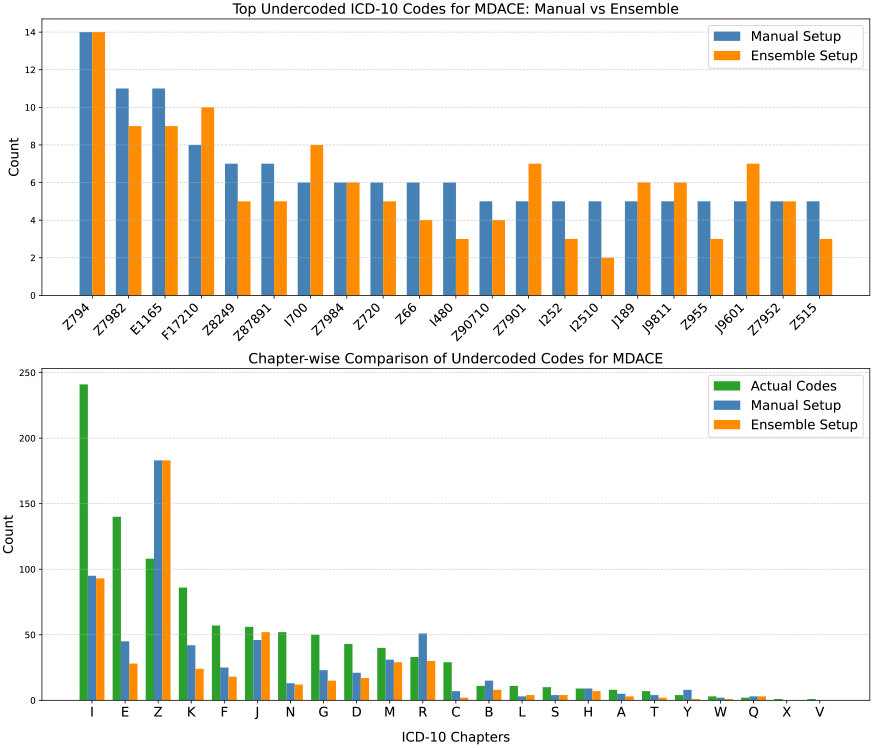
MDACE Undercoding: (Top) the ensemble approach and manual review identify similar undercoded codes; (Bottom) undercoding is more common in high-frequency chapters.

On the other hand, undercoding in CodiEsp was minimal, with an average rate of only **5.3%**, possibly due to the fact that the texts in CodiEsp (i.e., short death reports) are often densely coded. Again, the limited undercoding observed was mostly concentrated in Z-codes and followed a similar frequency-driven pattern as seen in MDACE. Given its low incidence, we did not conduct a manual review of undercoding on CodiEsp.

### C. Error Detection Findings

We found an overall error rate of **10.35%** for MDACE using the ensemble setup. When we compared the predicted codes against the original annotations during manual annotation, the error rate was **9.80%**. The errors were distributed across eight distinct error types, as detailed in Table I. In CodiEsp, the error rate was much higher than in MDACE, about **22.26%** in the ensemble setup and rising to **29.74%** based on manual review. Among the 250 manually reviewed clinical notes, the original annotations included a total of 2,841 codes. Comparison with the predicted codes revealed 845 error cases.

MDACE predominantly exhibited errors related to Clinical Detail Mismatch, followed by General Overcoding. These involved cases where *more specific or clinically accurate alternatives* to the original codes should have been assigned and *codes broader than warranted* should have been avoided respectively. Instances of assigning codes based on negative results (1.46% for False Positive) or codes lacking supporting evidence (5.11% for No Evidence) were rare in MDACE, likely due to the presence of human-annotated evidence spans.

CodiEsp, in contrast, showed overcoding as a dominant issue. The most error-prone categories are General Overcoding (25.6%) and Signs/Symptoms (24.26%). Unlike MDACE too, in CodiEsp we identified a significant portion of clinical notes in which diagnoses explicitly ruled out (i.e., documented as negative findings) were nonetheless coded (20.95% for False Positives), thereby violating official ICD-10 coding guidelines. As illustrated in Fig. 6 and Fig. 7 respectively, we noted that the error codes in both MDACE and CodiEsp followed the same distribution pattern as the undercoded codes and the most frequent codes and chapters were also the most error-prone. Manual and ensemble analyses produced comparable results, which underscores its high potential for clinical coding audit.

**Fig 6.**
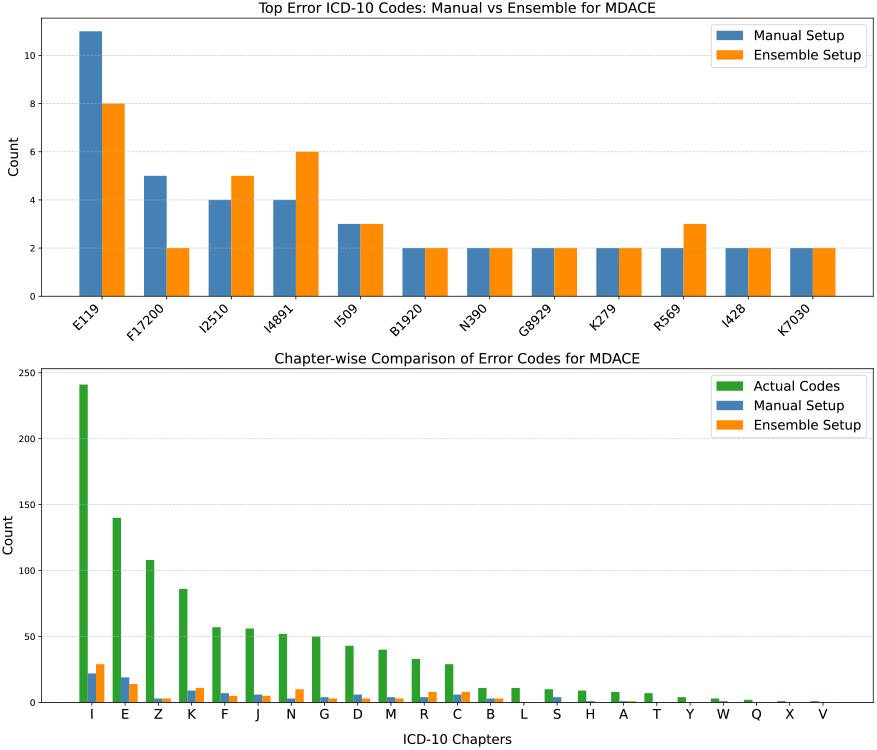
Manual and ensemble methods identify similar errors on MDACE: (Top) Most frequent erroneous codes by the manual method; (Bottom) Coding errors are more common in high-frequency chapters.

**Fig 7.**
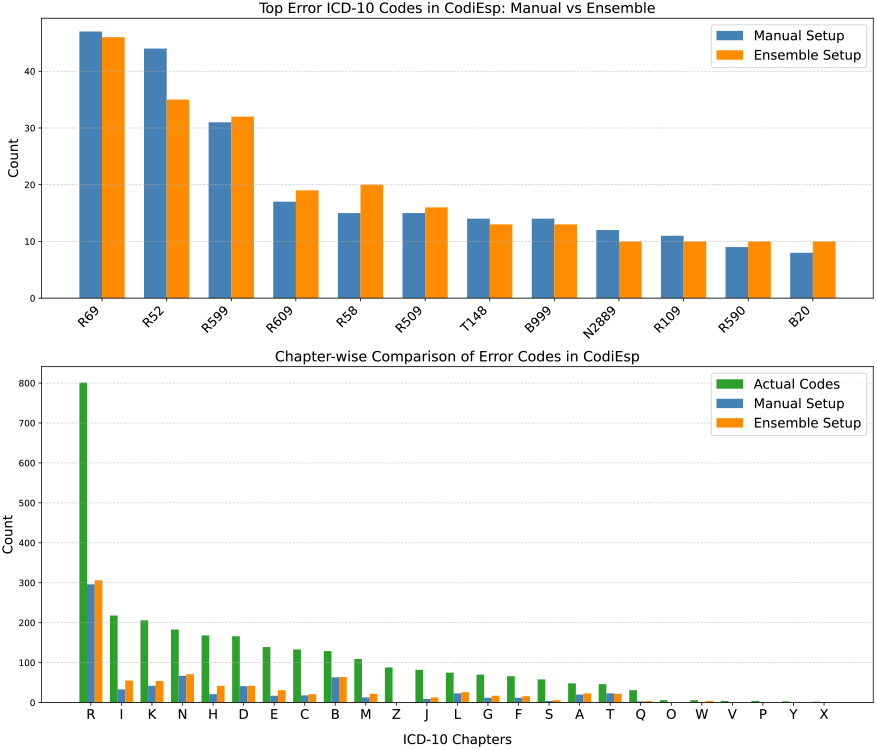
Manual and ensemble methods detect similar errors on CodiEsp: (Top) Most frequent erroneous codes by the manual method; (Bottom) Coding errors are more common in high-frequency chapters.

### D. Correcting Errors Benefits Automated Clinical Coding

The identified coding errors were substituted by the correct replacements to form a gold-standard dataset based on (3). To demonstrate the impact of code quality, we evaluated the baseline ACC generated by our pipeline were incorporated into the revised models on both the original and revised gold-standard sets. The impact differed across the two datasets. In overall speaking, improving the quality of code dataset could lead to ACC models of notably improved performances.

For MDACE, we observed a clear improvement across all evaluation metrics when tested on the corrected dataset (Table II where performance improvements are indicated by upward arrows). Since the models were trained on MIMIC-IV, they seem to generalise well to MDACE. The improvements were relatively small because only coding errors were handled, the percentage of which is relatively small too, only around 10%. If undercoding were also considered, we could see much more visible performance gains.

**TABLE II.** Impact of Correcting Errors on MDACE.

| Model | Prompt | Micro |  |  | Macro |  |  |
| --- | --- | --- | --- | --- | --- | --- | --- |
|  |  | Prec | Rec | F1 | Prec | Rec | F1 |
| CAML | MDACE | 0.332 | 0.491 | 0.396 | 0.163 | 0.205 | 0.166 |
| CAML | MDACE+ | 0.337<sup>↑</sup> | 0.521<sup>↑</sup> | 0.409<sup>↑</sup> | 0.172<sup>↑</sup> | 0.221<sup>↑</sup> | 0.178<sup>↑</sup> |
| MultiResCNN | MDACE | 0.689 | 0.354 | 0.468 | 0.192 | 0.191 | 0.183 |
| MultiResCNN | MDACE+ | 0.690<sup>↑</sup> | 0.372<sup>↑</sup> | 0.483<sup>↑</sup> | 0.198<sup>↑</sup> | 0.197<sup>↑</sup> | 0.188<sup>↑</sup> |
| LAAT | MDACE | 0.464 | 0.413 | 0.437 | 0.229 | 0.235 | 0.219 |
| LAAT | MDACE+ | 0.473<sup>↑</sup> | 0.441<sup>↑</sup> | 0.457<sup>↑</sup> | 0.248<sup>↑</sup> | 0.258<sup>↑</sup> | 0.240<sup>↑</sup> |
| PLM-ICD | MDACE | 0.635 | 0.603 | 0.618 | 0.341 | 0.353 | 0.335 |
| PLM-ICD | MDACE+ | 0.639<sup>↑</sup> | 0.639<sup>↑</sup> | 0.639<sup>↑</sup> | 0.347<sup>↑</sup> | 0.368<sup>↑</sup> | 0.346<sup>↑</sup> |

For CodiEsp, the improvements were less pronounced (Table III) despite a higher error rate. This is partly because the training data combined both MIMIC-IV and CodiEsp, which introduced inconsistency. Removing erroneous codes improved recall across all models, but precision dropped, reflecting noise introduced by incorrect labels in the training set.

**TABLE III.** Impact of Correcting Errors on CodiEsp.

| Model | Prompt | Micro |  |  | Macro |  |  |
| --- | --- | --- | --- | --- | --- | --- | --- |
|  |  | Prec | Rec | F1 | Prec | Rec | F1 |
| CAML | CodiEsp | 0.425 | 0.387 | 0.405 | 0.225 | 0.218 | 0.207 |
| CAML | CodiEsp+ | 0.365 | 0.409<sup>↑</sup> | 0.385 | 0.215 | 0.224<sup>↑</sup> | 0.209<sup>↑</sup> |
| MultiResCNN | CodiEsp | 0.347 | 0.233 | 0.278 | 0.120 | 0.123 | 0.111 |
| MultiResCNN | CodiEsp+ | 0.308 | 0.266<sup>↑</sup> | 0.285<sup>↑</sup> | 0.118 | 0.133<sup>↑</sup> | 0.115<sup>↑</sup> |
| LAAT | CodiEsp | 0.383 | 0.493 | 0.431 | 0.216 | 0.254 | 0.218 |
| LAAT | CodiEsp+ | 0.316 | 0.504<sup>↑</sup> | 0.389<sup>↑</sup> | 0.204 | 0.257<sup>↑</sup> | 0.212 |
| PLM-ICD | CodiEsp | 0.535 | 0.400 | 0.457 | 0.251 | 0.239 | 0.227 |
| PLM-ICD | CodiEsp+ | 0.479 | 0.449<sup>↑</sup> | 0.463<sup>↑</sup> | 0.240 | 0.248<sup>↑</sup> | 0.229<sup>↑</sup> |

### E. Performance of the Code Prediction Pipeline

#### 1) LLMs Can Extract Medical Evidences That Support ICD-10 Coding

On MDACE, all extractors obtained a recall of **over 95%** (Table IV) with DeepSeek-R1 giving the best recall. LLMs often extracted relevant but unannotated text. This leads to unfairly low precision as evidence text spans valid for coding are largely under-annotated in MDACE. For example, several pieces of semantically equivalent evidence in different places of a clinical note may correspond to an assigned code, but only one is annotated on most occasions. Our method significantly outperformed the recent LLM-based methods, particularly in recall, such as CLH-large [57] (62% recall) and Jiang et al.’s initial results in [62] (65.9% by GPT.3.5, 69.5% by finetuned BioBERT). Evidence extractio n was not evaluated on CodiEsp because we decided to omit is as described in Sect. IV.B.

**TABLE IV.** Performance of Evidence Extractors on MDACE.

| Model | Precision | Recall | F1 |
| --- | --- | --- | --- |
| GPT-4.1 | 0.111 | 0.959 | 0.199 |
| Gemini-1.5-Pro | 0.085 | 0.962 | 0.148 |
| Gemini-2.0-Flash | 0.092 | 0.959 | 0.168 |
| DeepSeek-R1 | 0.088 | 0.966 | 0.161 |
| GPT-3.5 zero-shot [12] | 0.248 | 0.659 | 0.361 |
| BioBERT finetuned [12] | 0.535 | 0.695 | 0.604 |
| CLH-large [41] | 0.06 | 0.62 | -- |

**TABLE V.**
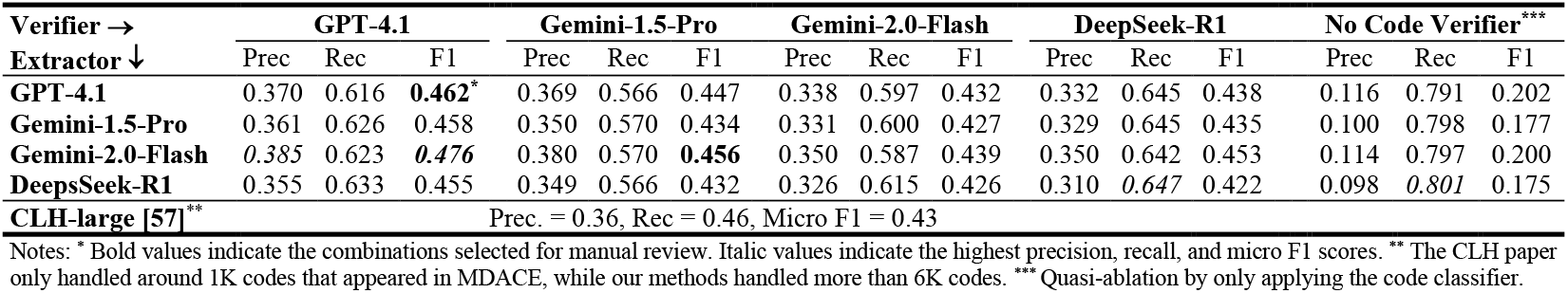
Performance of different verifier–extractor combinations on MDACE.

| Verifier →<br>Extractor ↓ | GPT-4.1 |  |  | Gemini-1.5-Pro |  |  | Gemini-2.0-Flash |  |  | DeepSeek-R1 |  |  | No Code Verifier*** |  |  |
| --- | --- | --- | --- | --- | --- | --- | --- | --- | --- | --- | --- | --- | --- | --- | --- |
|  | Prec | Rec | F1 | Prec | Rec | F1 | Prec | Rec | F1 | Prec | Rec | F1 | Prec | Rec | F1 |
| GPT-4.1 | 0.370 | 0.616 | **0.462\*** | 0.369 | 0.566 | 0.447 | 0.338 | 0.597 | 0.432 | 0.332 | 0.645 | 0.438 | 0.116 | 0.791 | 0.202 |
| Gemini-1.5-Pro | 0.361 | 0.626 | 0.458 | 0.350 | 0.570 | 0.434 | 0.331 | 0.600 | 0.427 | 0.329 | 0.645 | 0.435 | 0.100 | 0.798 | 0.177 |
| Gemini-2.0-Flash | 0.385 | 0.623 | **0.476** | 0.380 | 0.570 | **0.456** | 0.350 | 0.587 | 0.439 | 0.350 | 0.642 | 0.453 | 0.114 | 0.797 | 0.200 |
| DeepSeek-R1 | 0.355 | 0.633 | 0.455 | 0.349 | 0.566 | 0.432 | 0.326 | 0.615 | 0.426 | 0.310 | *0.647* | 0.422 | 0.098 | *0.801* | 0.175 |
| CLH-large [57]** | Prec. = 0.36, Rec = 0.46, Micro F1 = 0.43 |  |  |  |  |  |  |  |  |  |  |  |  |  |  |

Notes: \* Bold values indicate the combinations selected for manual review. Italic values indicate the highest precision, recall, and micro F1 scores. \*\* The CLH paper only handled around 1K codes that appeared in MDACE, while our methods handled more than 6K codes. \*\*\* Quasi-ablation by only applying the code classifier.

#### 2) The Code Prediction Pipeline Is a Potential Framework for LLM-based ACC

The match rate between silver evidence and human-annotated evidence on MDACE was **84.7%** (*n* = 1), which included exact match (54.8%) and extended matches that subsume the annotated evidence (30.9%). This highlights the quality of silver evidence and its suitability for training the code classifier. The idea of silver evidence was further justified in additional experiments in Appendix. The code classifier that was trained on silver evidence (*n* = 1) achieved a high accuracy of **82.47%** on MDACE-labeled evidence. Interestingly, the codes with the least accuracies were also among the most error-prone codes identified in Sect. VI.C, so the real accuracy is expected to be much higher, which justifies the suitability of the multiclass classifier as code predictor.

Table V shows the performances of all extractor-verifier combinations on MDACE. To increase the recall, a threshold of 0.1 was applied to the posterior probability of the code classifier. Our coding pipeline outperformed the SoTA CLH-large model (which only worked on a 1K label space). GPT-4.1 consistently achieved the highest F1 scores as a verifier across combinations. When Deepseek-R1 served as the verifier we obtained the highest recall, while Gemini-2.0-flash as the extractor achieved the highest precision. Table VI demonstrates the performances of the code verifier on CodiEsp, which significantly outperformed evidence-based verification in [29]. In [29], PLM-ICD was used for generating an initial set of codes, but PLM-ICD does not generalise well to CodiEsp. By contrast, our code classifier was trained on sentence-/phrase-level silver evidence, it proved to generalise better to CodiEsp.

**TABLE VI.** Performances on Code Verifiers on CodiEsp.

| Model (as Code Verifier) | Precision | Recall | F1 |
| --- | --- | --- | --- |
| GPT-4.1 | 0.753 | 0.384 | 0.509 |
| Gemini-1.5-Pro | 0.664 | *0.311* | 0.423 |
| Gemini-2.0-Flash | 0.606 | 0.482 | *0.537* |
| DeepSeek-R1 | 0.579 | 0.380 | 0.459 |
| Evidence-based verification (GPT-4o) [54]* | 0.306 | 0.302 | 0.304 |

\* Best reported performance in [54].

We also evaluated on MIMIC-IV-ICD10-Mini, a subset of 250 random MIMIC-IV notes used in [54] (Table VII). As in [54], we used GPT-4o to replace GPT-4.1. Again, GPT-4o consistently achieved better precision as an extractor and DeepSeek as a verifier better in recall. Our method slightly underperformed but were close to that of [54]. Our recall was lower, possibly due to some codes being unpredictable as codes appearing less than 5 times were excluded in the current study. In contrast, the sampled codes for verification had a high recall (0.7854 in Table II in [54]), which resulted in a better F1.

**TABLE VII.** Performances of the Pipeline on MIMIC-IV-ICD10-Mini.

| Verifier →<br>Extractor ↓ | GPT-4o |  |  | DeepSeek-R1 |  |  |
| --- | --- | --- | --- | --- | --- | --- |
|  | Prec | Rec | F1 | Prec | Rec | F1 |
| GPT-4o | 0.375 | 0.463 | *0.417* | 0.361 | 0.500 | *0.420* |
| DeepSeek-R1 | 0.347 | 0.464 | 0.399 | 0.316 | 0.502 | 0.373 |
| GPT-4o - Extractor Only | Prec = 0.136, Rec = 0.599, F1 = 0.221 |  |  |  |  |  |
| DeepSeek-R1 - Extractor Only | Prec = 0.118, Rec = 0.631, F1 = 0.201 |  |  |  |  |  |
| Evidence-based verification [54] | Prec = 0.339, Rec = **0.640**, F1 = **0.443** |  |  |  |  |  |

**TABLE VIII.** F1 Scores of PLM-ICD Using Silver Evidences on MDACE.

| Train ↓ Test → | Top 1 | Top 3 | Top 5 | Top 9 | MIMIC-IV |
| --- | --- | --- | --- | --- | --- |
| Top 1 | **0.688** | 0.604 | 0.543 | 0.497 | 0.427 |
| Top 3 | 0.611 | **0.725** | 0.697 | 0.637 | 0.485 |
| Top 5 | 0.591 | 0.680 | **0.731** | 0.696 | 0.513 |
| Top 9 | 0.575 | 0.616 | 0.699 | **0.723** | 0.531 |
| MIMIC-IV | 0.225 | 0.534 | 0.541 | 0.532 | 0.594 |

#### 3) Quasi-Ablation Studies

The “No Code Verifier” column in Table V and “Extractor Only” row in Table VII show the results of the quasi-ablation studies. Without code verification, better recalls were achieved at the sacrifice of precision. The best recall on MDACE was quite high (0.8 by DeepSeek-R1 and over 0.79 by other models). Recall dropped by 10-15% on MIMIC-IV-ICD10-Mini, resulting in a lower F1. It is a key success factor to attain high recall before code verification.

### F. Potential for Clinical Coding Audit

To validate the efficacy of the LLM ensemble for clinical coding audit, we benchmarked it against a revised gold standard of MDACE that is derived from the manual review, which is defined as: Revised Gold Codes = Correct Codes (predicted by the LLM ensemble) − Incorrect Codes (identified by human reviewer) + Undercoded Codes (annotated by human reviewer). We performed a sensitivity analysis of the LLM ensemble’s performance by sweeping the voting threshold from 8 to 16 and calculating the resulting precision, recall, and F1 scores against these manually validated gold codes.

The LLM ensemble method demonstrated high performance across all tested thresholds on MDACE, all achieving micro F1 above 0.74 (Fig. 8). As shown in the performance curves, the optimal balance between precision and recall occurred at the threshold of 12 (Max F1: 0.7912), which matched the strict two-thirds majority threshold used in (2). These results indicate that a majority-voting ensemble based on LLMs can approximate human level accuracy in clinical coding. This strongly supports our primary hypothesis: ensemble analysis is a viable, scalable alternative to manual clinical coding auditing, particularly when optimized at a two-thirds majority threshold.

**Fig 8.**
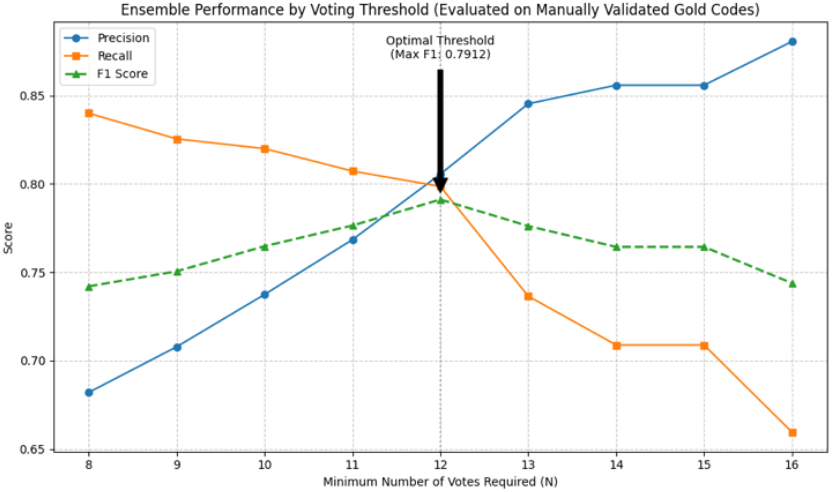
Performances of LLM ensembles on MDACE based on different thresholds for majority-voting according to (2).

### G. Discussions, Limitations and Future Work

Our explorative study echoed the findings by prior studies that coding quality issues are prevalent and significant. Most prior manual reviews treated undercoding as one type of coding error [16-25]. Similar to prior studies by pure manual review [16], we found undercoding to be the most prevailing code quality issue in hospital discharge summaries and, when compared with them, our undercoding rate seems to be among the highest, like [18]. This high undercoding rate on one hand risks invalidating the findings of population health research which relies on ICD codes to identify samples of a particular disease and on the other hand becomes a major source of renumeration loss of health care providers. Thus, our coding pipeline has significant values in both senses. However, the current study did not cover how and how much undercoding in our datasets impacts financial renumeration. In contrast, other types of error appear much less frequently in hospital discharge summaries than in death certificates (CodiEsp in Spanish) as the latter tends to be heavily overcoded.

One of the most important conjectures of the current study is that evidence-based interpretable coding is the right direction of future work. Experiments showed that even silver evidence has high value for training an accurate code predictor. If we could train a code predictor on evidence texts that are more codable or better explain the link to the correct codes, we should expect the code predictor to generalise better to different genres of clinical texts with more stable performance, as demonstrated by the experimental results on CodiEsp. However, our current implementation has several limitations, which require further research. Firstly, although our definition of rare code is much stricter (≤ 5 times) than most studies (≤ 10, 20 or 50 times), their exclusion resulted in an inherent loss of prediction capability for about 20,000 codes that appear, although very infrequently, in MIMIC-IV. In addition, our evidence extractor is suboptimal. The current implementation can find high-quality candidate evidence but cannot handle codes that require multiple pieces of evidence. Nevertheless, the current study has made the first step towards large-scale mining of coding evidence/rationale. Few studies have touched similar things [44, 46].

Our work advocates for evidence-based clinical coding also because it holds great value in uncovering coding errors. The proposed transparent pipeline enables coders to clearly trace which evidence leads to which code, and whether that code is valid or not. This approach allowed us to pinpoint the causes of errors and address them systematically, as demonstrated by the manual reviews of LLM-based suggestions for potential coding errors in this study. This method also shows promise for adaptation into practical code recommendation systems.

To achieve evidence-based clinical coding, it is of utmost importance to extract codable evidence texts. We believe it is not an impossible task. The current study has demonstrated, at least partially, that LLMs can identify evidence texts that are annotated by human coders. The recalls on MDACE were close to perfect though the precision were low compared to finetuned models. The experiments on silver evidence demonstrated that the problem is not to extract multiples pieces of evidence texts for each code, as PLM-ICD models trained using top-3, top-5, top-7, top-9 silver evidence all achieved over 20% relative coding performance improvements, but to extract only pertinent evidence and reduce noise that is introduced by LLM-based evidence extractor due to its tendency of over-extraction [11, 62]. We envision a further evidence verification step by either exploiting LLMs’ reasoning capabilities or building a bespoke evidence verifier targeted for clinical coding

With regards to the interpretable code prediction pipeline, we believe attaining high recall of the code predictor is one of the key success factors. The evidence-based code verifier in [54] relied on deep learning models to suggest a few dozens of codes and perform evidence extraction and code verification in one prompt. Our approach (code classifier) obtained high recall on MDACE and thus a good final performance. However, there was an obvious tradeoff between precision and recall. When the code verifier was applied, precision got improved but still not satisfactory enough when compared to deep learning models like PLM-ICD, and recall dropped often significantly. The same phenomenon also appeared in [54] and other LLM-based coding methods [57]. Improving code verification performance is an important future research direction for us.

Overlooking the quality issues of clinical coding dataset makes it difficult to realise the true potentials of machine learning algorithms when they are trained on subpar or flawed datasets. Coding audit and miscoding correction are valuable. Although our pipeline did not achieve comparable performance to deep learning-based models like PLM-ICD as existing LLM-based methods don’t either (despite beating some most recent approaches on MDACE and/or CodiEsp [54, 57]), it shows high values in serving as either an assistive coding audit tool or a complimentary code recommendation tool. Particularly, nearly one out of every two undercoded codes that were suggested by the ensemble setup of our approach were truly undercoded, which may also assist coding auditors in locating and resolving many coding errors that constituted over 10% of all assigned codes in MIMIC-style discharge summaries and nearly 30% in Spanish death certificates. In addition, our results also validated the use of LLM-based ensemble methods for both undercoding detection and code verification. Across all experimental setups, we observed strong alignment between ensemble outcomes and the results of manual review, confirming the value of ensemble approaches in supporting reliable, scalable coding verification, which could serve as a powerful tool for clinical coding audit.

## VII. Conclusion

This paper proposes an interpretable clinical coding pipeline and performed an in-depth analysis of the code quality issues, including undercoding and other coding errors, of two popular datasets for evidence-based clinical coding. In long clinical notes like hospital discharge summaries in MIMIC datasets, the undercoding ratio can be very high, over 70% in our explorative study. In shorter and denser clinical notes like death certificates in CodiEsp, coding errors could be high, close to 30% in our analysis and overcoding could be a significant issue. Correcting coding errors can bring consistent performance improvements to state-of-the-art automated coding algorithms. The results underscore the necessity of shifting research focus from model-centric to data-centric solutions in automated clinical coding. The proposed pipeline is seen as a stone that kills two birds. It is a highly potential framework for evidence-based coding as well as an effective assistive tool for clinical coding audit. As LLMs have proved to be remarkably successful in extracting all human-annotated evidence, the current study envisions several key aspects for achieving a satisfactory LLM-based coding solution, including improving evidence verification, rare code prediction and code verification, as well as attaining high recall of code prediction before code verification.

## Data Availability

All data produced in the present work are contained in the manuscript.

## Appendix

Table VIII shows that the coding performance of a state-of-the-art deep learning model PLM-ICD were significantly improved by training and evaluating on top-*n* silver evidence texts that were extracted using the method described in Sect. III.C.1. When *n* = 3,5,9, the relative performance improvements in F1 were as huge as 21-23%.

